# Genetic architecture of the limbic white matter microstructure in aging and Alzheimer’s Disease

**DOI:** 10.1101/2025.05.19.25327915

**Authors:** Anna Lorenz, Aditi Sathe, Yisu Yang, Alaina Durant, Yiyang Wu, Michael E. Kim, Chenyu Gao, Nancy R. Newlin, Karthik Ramadass, Praitayini Kanakaraj, Nazirah Mohd Khairi, Zhiyuan Li, Tianyuan Yao, Yuankai Huo, Logan Dumitrescu, Niranjana Shashikumar, Kimberly R. Pechman, Shannon L. Risacher, Lori L. Beason-Held, Yang An, Konstantinos Arfanakis, Guray Erus, Christos Davatzikos, Mohamad Habes, Di Wang, Duygu Tosun, Arthur W. Toga, Paul M. Thompson, Elizabeth C. Mormino, Panpan Zhang, Kurt Schilling, Alzheimer’s Disease Neuroimaging Initiative (ADNI), The BIOCARD Study Team, The Alzheimer’s Disease Sequencing Project (ADSP), Marilyn Albert, Walter Kukull, Sarah A. Biber, Bennett A. Landman, Sterling C. Johnson, Barbara Bendlin, Julie Schneider, David A. Bennett, Angela L. Jefferson, Susan M. Resnick, Andrew J. Saykin, Timothy J. Hohman, Derek B. Archer

## Abstract

**Background:** Limbic white matter (WM) abnormalities are prevalent in aging and Alzheimer’s disease (AD), yet their underlying biological mechanisms remain unclear. This study aims to identify the genetic architecture of limbic WM microstructure in older adults by leveraging harmonized data from multiple cohorts, including those enriched for cognitively impaired individuals.

**Methods:** We analyzed diffusion MRI (dMRI) data from 2,614 non-Hispanic White older adults (mean age = 73.7 ± 9.8 years; 57% female; 26% cognitively impaired) across 7 harmonized aging cohorts. WM microstructure was assessed in 7 limbic tracts, including the cingulum, fornix, inferior longitudinal fasciculus (ILF), uncinate fasciculus (UF), and transcallosal tracts of the inferior, middle, and superior temporal gyri (ITG, MTG, STG) using advanced diffusion MRI metrics corrected for free-water (FW): fractional anisotropy (FA_FWcorr_), axial diffusivity (AxD_FWcorr_), mean diffusivity (MD_FWcorr_), radial diffusivity (RD_FWcorr_). We performed heritability estimations, genome-wide association studies (GWAS) and post-GWAS analyses (genetic covariance, gene-level and pathway analysis, transcriptome-wide association [TWAS] studies). The AD relevance of the discovered variants was explored using bulk RNA-seq data from caudate, dorsolateral prefrontal, and posterior cingulate cortex human brain tissues.

**Results:** Limbic WM microstructure demonstrated significant heritability (estimates between 0.26 and 0.60, *p*_*FDR*_ < 0.05 for 15 of 35 tract-by-microstructure combinations). GWAS identified 6 genome-wide significant loci (*p* < 5.0×10^−8^) associated with WM microstructure. Notably, for MTG RD_FWcorr_, we identified a locus on chromosome 18 (lead SNP: rs12959877) comprising 38 SNPs that are eQTLs for *CDH19*, a gene involved in cell adhesion and highly expressed in oligodendrocytes. Other significant associations involved SNPs near *KC6, SENP5, RORA, FAM107B*, and *MIR548A1*. Bulk RNA-seq analyses revealed that brain tissue expression of *RORA, FAM107B*, and *KC6* was significantly associated with cognitive decline and several AD pathologies (*p*_FDR_ < 0.05). Post-GWAS analyses identified the genes *SERPINA12* and *DNAJB14*, and highlighted the involvement of insulin signaling, immune response, and neurotrophic pathways. Genetic covariance analyses indicated shared genetic architecture between limbic WM and lipid profiles (e.g., HDL cholesterol), cardiovascular traits, and neurological conditions (e.g., multiple sclerosis) (*p*_FDR_ < 0.05).

**Conclusion:** This multi-cohort imaging genetics study identified several novel genes and biological pathways associated with limbic WM microstructure in an aging population enriched for cognitive impairment. The association of several identified genes with cognitive decline and AD pathology underscores their AD relevance. Our findings further suggest that the genetic underpinnings of limbic WM microstructure are linked to vascular health and inflammation, highlighting these pathways as promising avenues for future AD-related therapeutic development.

## Introduction

The integrity of nerve fibers and the myelination of axons, which characterize white matter (WM) microstructure, are essential for efficient cognitive processing by enabling communication across different brain regions.^1,2^ However, WM undergoes substantial changes during aging and in neurodegenerative diseases such as Alzheimer’s disease (AD).^3^ While AD has typically been viewed as a disorder primarily affecting gray matter, emerging evidence highlights distinct WM abnormalities, including axonal loss, demyelination, and microglial activation, even in early disease stages.^4–6^ Our group and others have demonstrated that WM abnormalities, particularly within the limbic system, are prevalent across the AD diagnostic continuum,^7^ with accelerated WM degeneration in individuals exhibiting abnormal aging^8^ and carriers of the apolipoprotein (*APOE*) ε4 allele.^9^ These WM changes can manifest before the appearance of AD pathologies such as amyloid-β plaques and neurofibrillary tangles,^5^ are detectable up to 22 years before symptom onset,^10,11^ and contribute to cognitive decline independently of gray matter atrophy such as hippocampal volume loss.^12^

Diffusion MRI (dMRI) is a non-invasive technique that allows for the *in vivo* quantification of WM microstructure by measuring water diffusion.^13–16^ By estimating the magnitude and directionally of diffusivity within each voxel, the method can quantify several conventional metrics, including fractional anisotropy (FA_CONV_), axial diffusivity (AxD_CONV_), mean diffusivity (MD_CONV_), and radial diffusivity (RD_CONV_). FA_CONV_ reflects the directional dependence of water diffusion, with higher values typically indicating compact WM tracts. AxD_CONV_ measures diffusion along the primary axis of fibers, where lower values can indicate axonal damage. MD_CONV_ represents the average diffusion rate, providing a general tissue density and cellularity measure. RD_CONV_ measures diffusion perpendicular to the primary axis, with higher values often suggesting demyelination.^13–16^ These conventional metrics, however, derived from single-tensor models can be confounded by partial volume effects of brain tissue with extracellular free water (FW), such as cerebrospinal fluid. Advanced bi-tensor models address this by estimating and correcting these metrics for FW contributions, yielding more biologically precise metrics of WM microstructure (FA_FWcorr_, AxD_FWcorr_, RD_Fwcorr,_ MD_FWcorr_) that differentiate between extracellular (i.e., FW) and intracellular contribution to water diffusion.^17^ Prior studies suggest that these FW-corrected metrics offer a more comprehensive assessment of WM neurodegenerative patterns in AD.^7–9,12^

*In vivo* measurements of WM microstructure have enabled the examination of the underlying biological mechanisms involved and have thus far demonstrated that WM microstructure across the brain is significantly influenced by genetic factors. Twin studies have reported high heritability for dMRI metrics (up to 82%), indicating substantial genetic contributions to variations in WM microstructure.^18,19^ Large-scale genome-wide association studies (GWAS) primarily in cognitively unimpaired populations, such as the UK Biobank, have identified more than 100 genomic regions influencing WM microstructure and have revealed genetic links between WM integrity and various clinical traits such as stroke, major depressive disorder, schizophrenia, and attention deficit hyperactivity disorder.^20,21^ However, these studies may not fully capture genetic pathways specific to neurodegenerative processes due to the underrepresentation of cognitively impaired individuals. Investigations focused on AD have implicated candidate genes, such as *APOE* and *BIN1*, in WM alterations.^22–25^ Additionally, an increased genetic predisposition for AD correlates with reduced cingulum bundle FA_CONV_^26^ and several AD risk variants in *TMEM106B, PTK2B, WNT3*, and *APOE* are associated with WM microstructure in older adults.^27^ Nevertheless, a more comprehensive understanding of the genetic architecture and biological pathways influencing WM microstructural differences in older adults, particularly those at risk for or with AD, remains limited due to the scarcity of combined diffusion and genetic data in such cohorts. Therefore, examining genetic influences on WM microstructure in cohorts enriched for cognitive impairment is critical for advancing our understanding of neurodegenerative processes in aging and AD. Large-scale harmonization efforts, such as the Alzheimer’s Disease Sequencing Project Phenotype Harmonization Consortium (ADSP-PHC), in combination with advanced FW-corrected dMRI methods presents an unprecedented opportunity to investigate these imaging genetic associations.

This study assessed the role of genetic factors in WM microstructure alterations across 7 tracts of the limbic system, including the cingulum, fornix, inferior longitudinal fasciculus (ILF), uncinate fasciculus (UF), and the transcallosal tracts of the inferior temporal gyrus (ITG), the middle temporal gyrus (MTG), and the superior temporal gyrus (STG). These tracts are integral to memory function and show substantial alterations in aging and AD.^7,12,24^ By analyzing harmonized FW-corrected dMRI and genetic data from 2,614 older adults (26% cognitively impaired) across 7 established aging cohorts, we aim to identify novel genetic variants and biological pathways associated with limbic WM microstructure, ultimately providing insights into the mechanisms of WM degeneration in the context of aging and AD.

## Methods

### Participants

Diffusion MRI (dMRI) and genetic data were leveraged from 7 cohorts, including the Alzheimer’s Disease Neuroimaging Initiative (ADNI), Biomarkers of Cognitive Decline Among Normal Individuals (BIOCARD), Baltimore Longitudinal Study of Aging (BLSA), National Alzheimer’s Coordinating Center (NACC), Religious Orders Study and Rush Memory and Aging Project (ROSMAP), Vanderbilt Memory and Aging Project (VMAP), and the Wisconsin Registry for Alzheimer’s Prevention (WRAP). Data collection for ADNI (https://adni.loni.usc.edu) began in 2004 as a public-private partnership, gathering data from cognitively unimpaired individuals, those with mild cognitive impairment (MCI), and participants with dementia due to AD. The project’s goal was to investigate the associations between serial magnetic resonance imaging (MRI), positron emission tomography (PET), other biological markers, clinical and neuropsychological assessments, and the progression from MCI to early AD.^28^ ADNI-GO, ADNI2, and ADNI3 phases with dMRI data were included in this study. BIOCARD began data collection in 1995 at the National Institute of Mental Health (NIMH) and was later transferred to John Hopkins University with the goal of identifying preclinical biomarkers of cognitive decline and investigating variables that can predict future progression to AD among cognitively normal middle-aged individuals. Participants undergo extensive longitudinal evaluations, including neuropsychological testing, MRI scans, and collection of blood and cerebrospinal fluid samples.^29^ The BLSA neuroimaging substudy began data collection in 1994 and included dementia-free participants aged 55 to 85 years, with up to 10 years of prospective data collection at baseline. In 2009, the cohort was expanded to include BLSA participants aged 20 to 85 years as well as 3T MRI-based acquisition of dMRI data. NACC maintains a centralized data repository for the National Institute of Aging’s (NIA’s) Alzheimer’s Disease Research Centers (ADRC) Program, which currently includes 33 centers and 4 exploratory centers across the United States.^30–32^ The ROS cohort, which began in 1994, is an ongoing longitudinal study collecting clinical-pathological data on aging and AD. Participants in ROS are 65+ years-old Catholic nuns, priests, and brothers from various groups throughout the US.^33^ MAP is a longitudinal study that started in 1997, recruiting cognitively unimpaired participants.^33^ The VMAP cohort began longitudinal data collection in 2012 with the goal of understanding the relationship between vascular health and brain aging, enriched in older adults with MCI.^34^ The WRAP cohort began data collection in 2001, focusing on middle-aged adults with a parental history of AD. In 2004, the study expanded to include a control group of individuals without a parental history of AD. The primary objective of WRAP is to identify early biomarkers and risk factors for AD before clinical symptoms emerge.^35,36^

For all cohorts, participants provided written informed consent, and research was conducted in accordance with approved Institutional Review Board protocols. Secondary analysis of these data was approved by the Vanderbilt University Medical School Institutional Review Board. The study included 2,614 participants who were self-reported non-Hispanic White individuals aged between 50.1 and 100.9 years (mean = 73.7 ± 9.8 years) and 57% were female. **Table 1** provides an overview of data included for this study from the ADNI, BIOCARD, BLSA, NACC, ROSMAP, VMAP, and WRAP cohorts.

**TABLE 1.**
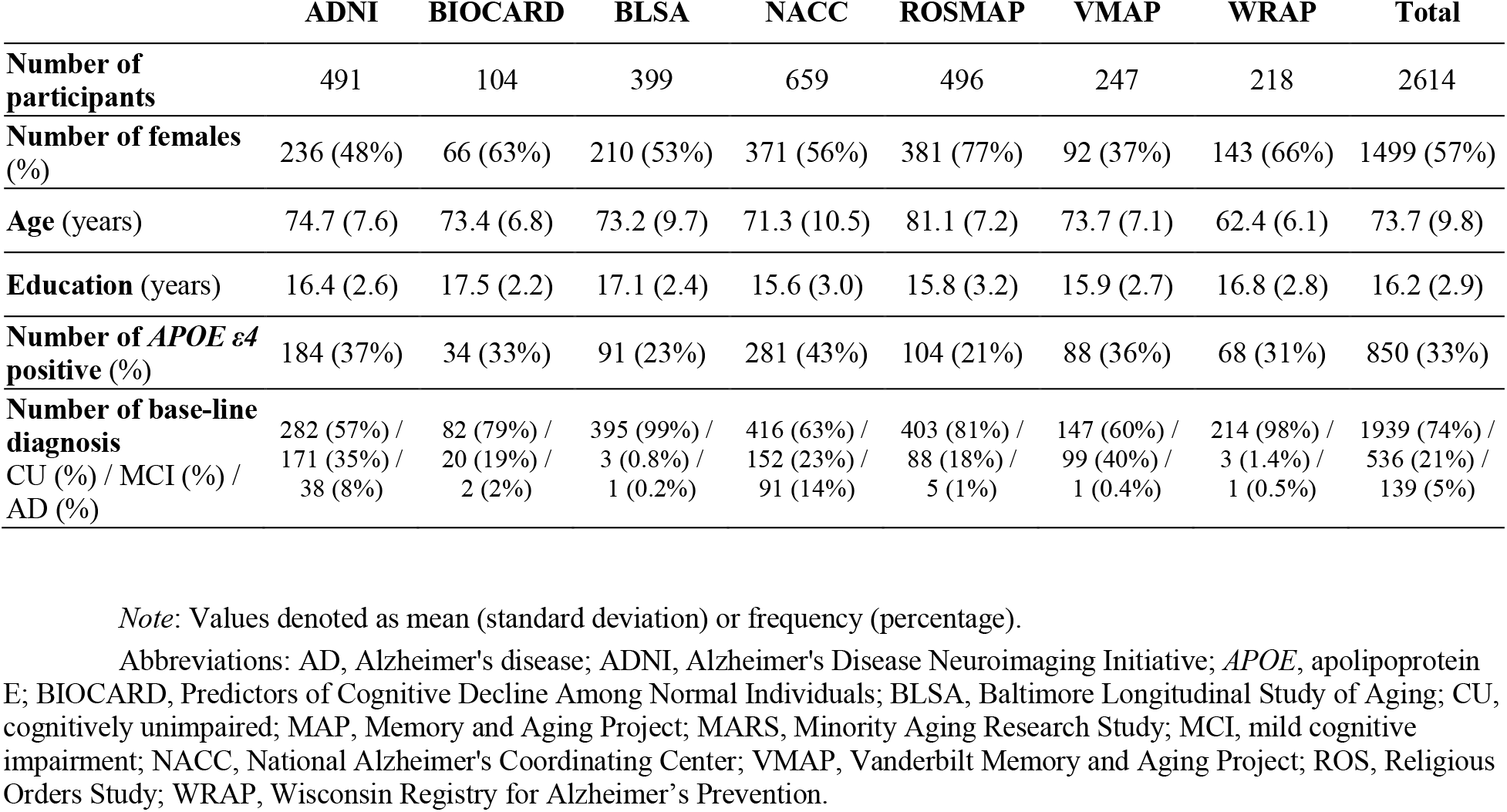
Participant characteristics by cohort

### Diffusion MRI acquisition and preprocessing

Across all cohorts, we had 78 different dMRI acquisition protocols – **Table S1** provides relevant parameters (e.g., number of directions, b-values, resolution). dMRI data from all cohorts were preprocessed using the *PreQual* pipeline, which addresses motion, distortions and eddy currents and performs slice-wise imputation.^37,38^ Imaging sessions were inspected manually by evaluating PDF reports outputted by the *PreQual* pipeline. DTIFIT was used for the remaining data to compute conventional dMRI metrics, including FA_CONV_, AxD_CONV_, MD_CONV_, and RD_CONV_. To account for FW content in each voxel, we calculated FW-corrected metrics, including FA_FWcorr_, AxD_FWcorr_, MD_FWcorr_, RD_FWcorr_ and FW.^17^ Symmetric normalization and linear interpolation was conducted using the Advanced Normalization Tools (ANTs) package to obtain a standard space representation of these maps by non-linearly registering the FA_CONV_ map to FMRIB58_FA atlas.^39^ The obtained warp from the registration was applied to all other microstructural maps. Individuals with significant age-regressed outliers (± 5 standard deviations) in WM tract microstructural values were excluded.^40^

### WM tractography templates

Tractography templates for this study were leveraged from existing resources^12,41,42^ and can be retrieved in a publicly available Zenodo repository.^43^ The focus for this study was on 7 WM tracts within the limbic system, specifically the cingulum, fornix, ILF, UF, as well as ITG, MTG, and STG transcallosal tracts.

### Data harmonization

For the dMRI data, a region of interest (ROI) approach was employed to calculate mean conventional and FW-corrected dMRI metrics for all tractography templates for each participant. These values were then harmonized using the *Longitudinal ComBat* package in R (version 4.1.0)^44^ applied to our entire in-house longitudinal dataset, including 5,144 participants across 10,346 imaging timepoints. This harmonization process utilized a batch variable which had varying levels of specificity depending on the cohort. Parameters used to create the batch variable can be found in **Table S2**.^7–9^ The batch variable accounted for various parameters across our cohorts, including scanner name, magnet strength, number of b-values/b-vectors, and resolution – this was optimized to reduce the number of batches while simultaneously accounting for parameters we most anticipated to account for between-batch heterogeneity. In total, we accounted for 34 unique batching levels. Additional covariates for the harmonization included mean-centered age, mean-centered age squared, education, race/ethnicity, cognitive status (cognitively unimpaired or cognitively impaired), *APOE*-ε4 positivity, *APOE*-ε2 positivity, and the interaction of mean-centered age and cognitive status.

The harmonized values were then scaled by their standard deviation. Next, the dataset was filtered to include only participants with genetic data and further narrowed to the baseline timepoint for cross-sectional analysis. We ultimately used FW-corrected dMRI measures across 7 limbic WM tracts (35 dMRI measures) for each participant.

### Genetic data quality control and imputation

Genetic data were collected with various genotyping arrays across and within cohorts (ADNI: Illumina Human610-Quad BeadChip, Illumina HumanOmniExpress BeadChip, Illumina Omni 2.5 M, Illumnia Global Screening Array v2; BIOCARD: Illumina OmniExpress; BLSA: Illumina HumanOmni2.5 BeadChip, Illumina HumanOmniExpress BeadChip; NACC: several different arrays were used to collect genetic data – acquisition of all genetic data is outlined on the NACC website [https://naccdata.org/nacc-collaborations/partnerships]; ROSMAP: Global Screening Array-24 v3.0 BeadChip, Affymetrix GeneChip 6.0, Illumina HumanOmniExpress; VMAP: Illumina HumanOmniExpress; WRAP: Illumina Human610, Illumina OmniExpress).

All genetic raw data underwent the same robust quality control and imputation pipelines.^45^ Variants that had a genotyping rate less than 95%, a minor allele frequency (MAF) less than 1% or deviated from Hardy-Weinberg Equilibrium (*p* < 1×10^−6^) were removed. In addition, participants were excluded if genotyping efficiency was poor (missing >1% of variants), if cryptic relatedness was present (PIHAT > 0.25) or if the reported and genotypic sex were not concordant. Imputation was performed on the University of Michigan Imputation Server using the TOPMed reference panel (hg38) with SHAPEIT phasing.^46^ Data were filtered to exclude variants with low imputation quality (R^2^ < 0.08), duplicated/multi-allelic variants, and MAF < 1%. Principal components analysis was conducted, and genetic ancestry outliers were excluded. 8 pairs of individuals were related across cohorts and were subsequently removed from the respective cohort with more individuals, namely BLSA, NACC, and ADNI.

### Statistical analyses

#### SNP-heritability tests

Single nucleotide polymorphism (SNP) heritability was estimated for each FW-corrected dMRI metric using Genome-Wide Complex Trait Analysis (GCTA). This method uses restricted maximum likelihood and genetic relatedness matrices to calculate heritability estimates.^47^ To account for multiple comparisons, results were adjusted across phenotypes using the False Discovery Rate (FDR) procedure.^48^

#### Genome-wide association testing and meta-analysis

GWAS in self-reported non-Hispanic White participants were conducted separately for each cohort (N = 7) for each dMRI metric (N = 5) of each of the limbic WM tracts (N = 7) using PLINK software (Version 1.9, https://www.cog-genomics.org/plink/1.9). All models included covariates for age, sex and the first three genetic ancestry principal components. Next, we performed a meta-analysis across cohorts for each dMRI metric using Genome-Wide Association Meta-Analysis (GWAMA).^49^ The significance threshold was set *a priori* to *p* < 5×10^−8^. We evaluated suggestive loci which had a *p* < 1×10^−5^. Reported genome-wide associations were filtered to include those present in at least 6 out of 7 cohorts.

#### eQTL analyses, replication, and databases

Expression quantitative trait locus (eQTL) analyses were conducted on variants reaching genome-wide significance using the Genotype-Tissue Expression (GTEx) portal (https://gtexportal.org). Furthermore, genome-wide significant variants were validated using the Oxford Brain Imaging Genetics Server (BIG40) (https://open.win.ox.ac.uk/ukbiobank/big40/)^50^ and ENIGMA-VIS (https://enigma-brain.org/).^51^ The genes identified were further explored using several databases including GeneCards (https://www.genecards.org), Agora (https://agora.adknowledgeportal.org), and Open Targets (https://www.opentargets.org). A literature search was also conducted using PubMed and Web of Science to provide further context and insights into the identified genes.

#### Association analysis with cognitive outcomes and AD pathologies using ROSMAP bulk RNA sequencing data

Processed bulk RNAseq data in the ROSMAP cohort from three brain tissues (**Table S5a**), including the dorsolateral prefrontal cortex (DLPFC), the posterior cingulate cortex (PCC), and the head of the caudate nucleus (CN),^52^ were used to fit linear regression models (for cross-sectional outcomes) and linear mixed-effect models (for longitudinal outcomes) to test associations between cognitive function and AD pathologies with expression of the genes closest to the identified genome-wide significant variants in the meta-analyzed GWAS.

Overall cognitive function was represented using a global cognitive score which was derived by converting raw scores from 19 cognitive tests to z-scores and subsequently averaged. For cross-sectional cognitive function, the global cognitive score at last visit before death was used. For the longitudinal model, cognitive trajectory was quantified in a mixed effects regression model. Cognitive trajectory was estimated as the individual-specific annual rate of change in global cognition. AD pathologies included amyloid-β load (immunohistochemistry staining), neurofibrillary tangles (silver staining), tau tangle (immunohistochemistry staining), and neuritic plaque pathology (silver staining) (https://www.radc.rush.edu/docs/var/overview.htm?category=Pathology).

For all cross-sectional outcomes, covariates included age at death, sex, postmortem interval (PMI), and interval between last visit and death. For the longitudinal models, in addition to the same set of covariates, time was modeled as the number of years between each visit and the final visit. To account for multiple comparisons, results were adjusted for each phenotype using the FDR correction.^48^

#### Gene- and pathway-level analysis

The Multimarker Analysis of GenoMic Annotation (MAGMA v1.09) software was used to conduct gene- and pathway-level analyses.^53^ Results were adjusted for each phenotype using FDR correction.^48^

#### Genetic covariance analysis and local genetic covariance analysis

The meta-analysis results for all dMRI metrics were used to perform genetic covariance analyses with the GWAS summary statistics of 65 complex traits (**Table S9a**) as well as 3,143 brain traits^54^ derived from the UK Biobank using the Genetic Covariance Analyzer (GNOVA) software.^55^ To further investigate local genetic covariance for significant associations, we used the Super Genetic Covariance Analyzer (SUPERGNOVA) software.^56^ To account for multiple comparisons, results were adjusted across dMRI metrics and traits using the FDR procedure.^48^

#### Transcriptome-wide association testing

By leveraging predictive models, transcriptome-wide association studies (TWAS) increase the power to detect associations with WM microstructure that might have been missed with the GWAS approach. S-PrediXcan was used to examine associations between genetically predicted gene expression and dMRI metrics, leveraging precomputed predictive gene expression models from all GTEx tissues.^57^ To account for multiple comparisons, results were adjusted for each phenotype across genes and tissues using the FDR procedure.^48^

## Results

### Heritability of limbic WM microstructure

15 of 35 FW-corrected dMRI metrics across all limbic tracts were heritable with SNP-heritability estimates ranging from 0.26 to 0.60 (*p*_*FDR*_ < 0.05, **Figure 1, Table S3**). Specifically, all dMRI metrics of the cingulum as well as 4 out of 5 metrics of the fornix and ILF showed significant heritability.

**FIGURE 1.**
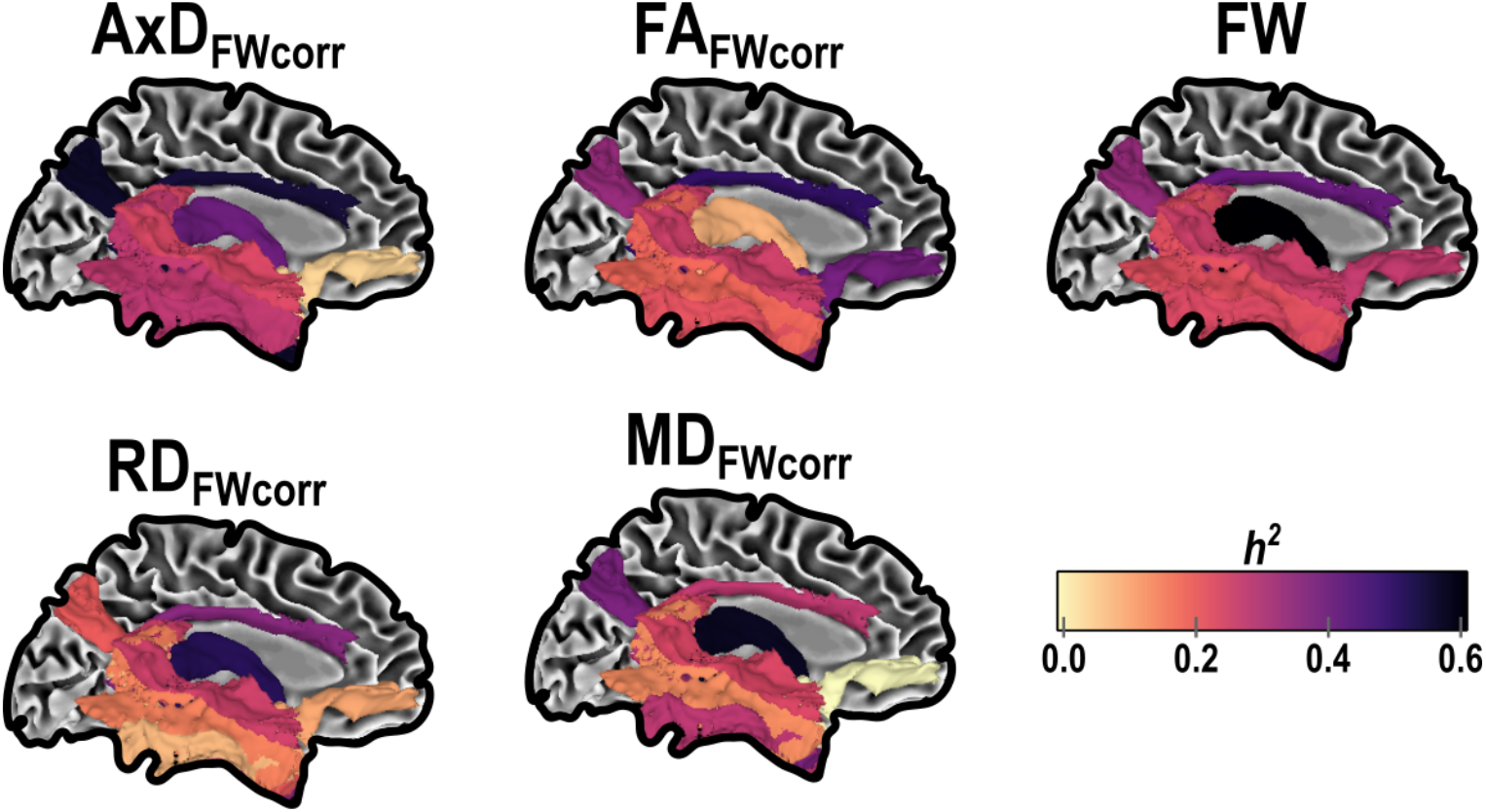
SNP-heritability estimates for limbic WM microstructure. SNP heritability of 35 FW-corrected dMRI metrics from 7 WM tracts in the limbic system. Abbreviations: AxD, axial diffusivity; dMRI, diffusion magnetic resonance imaging; FA, fractional anisotropy; FW_corr_, free water corrected; MD, mean diffusivity; RD, radial diffusivity; WM, white matter.

### Genome-wide significant signals in proximity to the genes *CDH19, KC6, SENP5, RORA, FAM107B*, and *MIR548A1* were associated with limbic WM microstructure

A cross-sectional GWAS for each dMRI metric of each tract was performed using linear regression models covarying for sex, age, and the first three ancestral principal components, followed by a meta-analysis across cohorts (**Table S4a** for meta-analysis summary statistics [*p* < 0.05]). **Figure 2** presents an ideogram illustrating the 500 most significant genomic signals based on smallest p-value associated with WM microstructure. In total, 6 genome-wide significant loci for different tract-by-microstructure combinations were discovered. We identified a locus with 38 genome-wide significant (*p* < 5×10^−8^) SNPs (**Figure 3**, lead SNP: rs12959877, EAF = 0.44, β = 0.002 ± 3.18×10^−4^, *p* = 5.78×10^−9^, intronic *CDH19*) for MTG RD_FWcorr_, with additional suggestive associations (*p* < 1×10^−5^) at this site for RD_FWcorr_ of STG and ITG. When evaluating eQTL (GTEx Portal) evidence, we found that all 38 SNPs were eQTLs for the gene *CDH19* in lung and spleen tissue (**Table S4b**). This gene is highly expressed in oligodendrocytes and functions as a calcium-dependent cell adhesion glycoprotein.^58^ Additional genome-wide significant signals were found in proximity to the genes *KC6, SENP5, RORA, FAM107B*, and *MIR548A1* (**Table 2**).

**TABLE 2.**
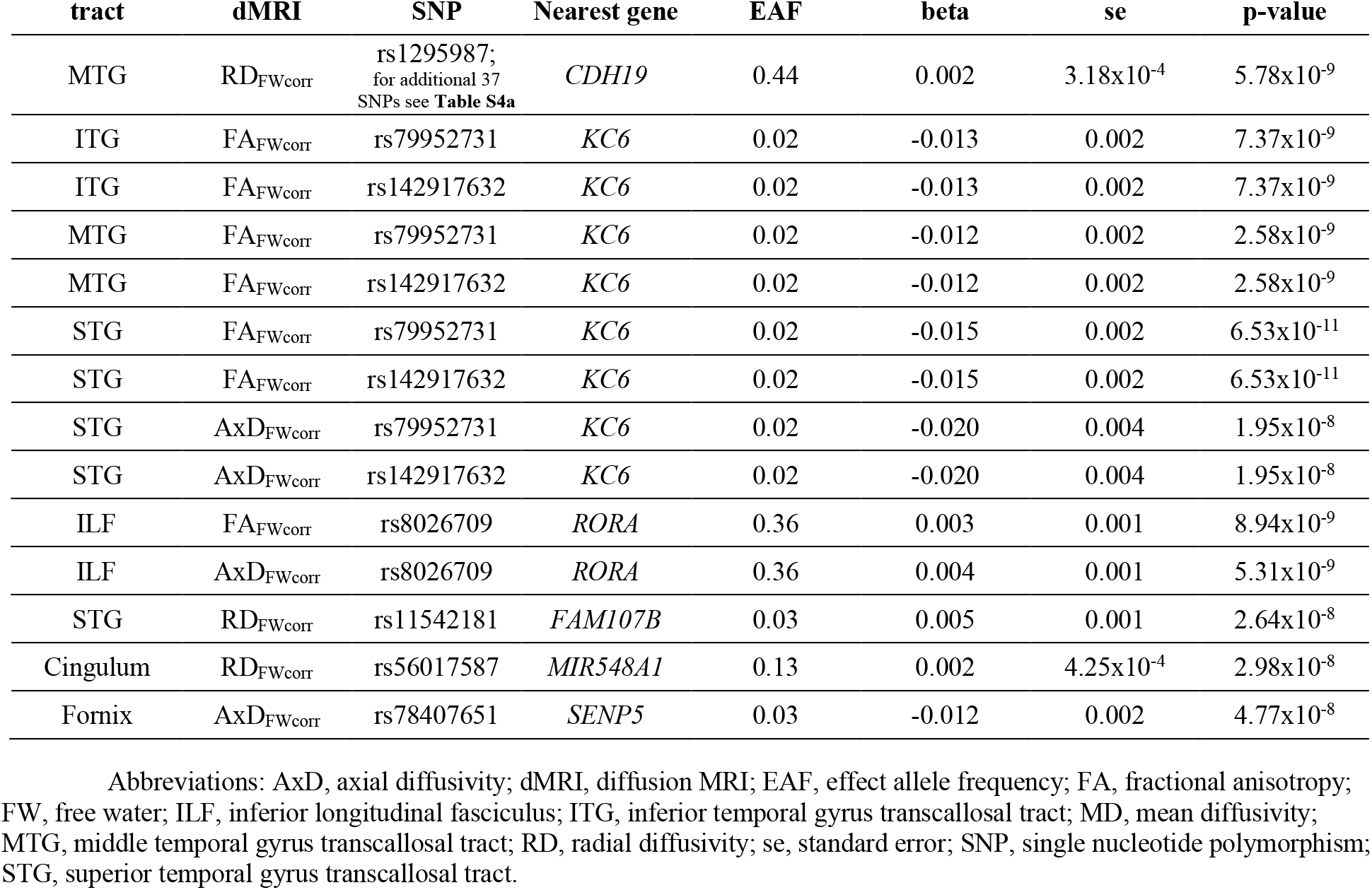
Statistics for genome-wide significant SNPs

**FIGURE 2.**
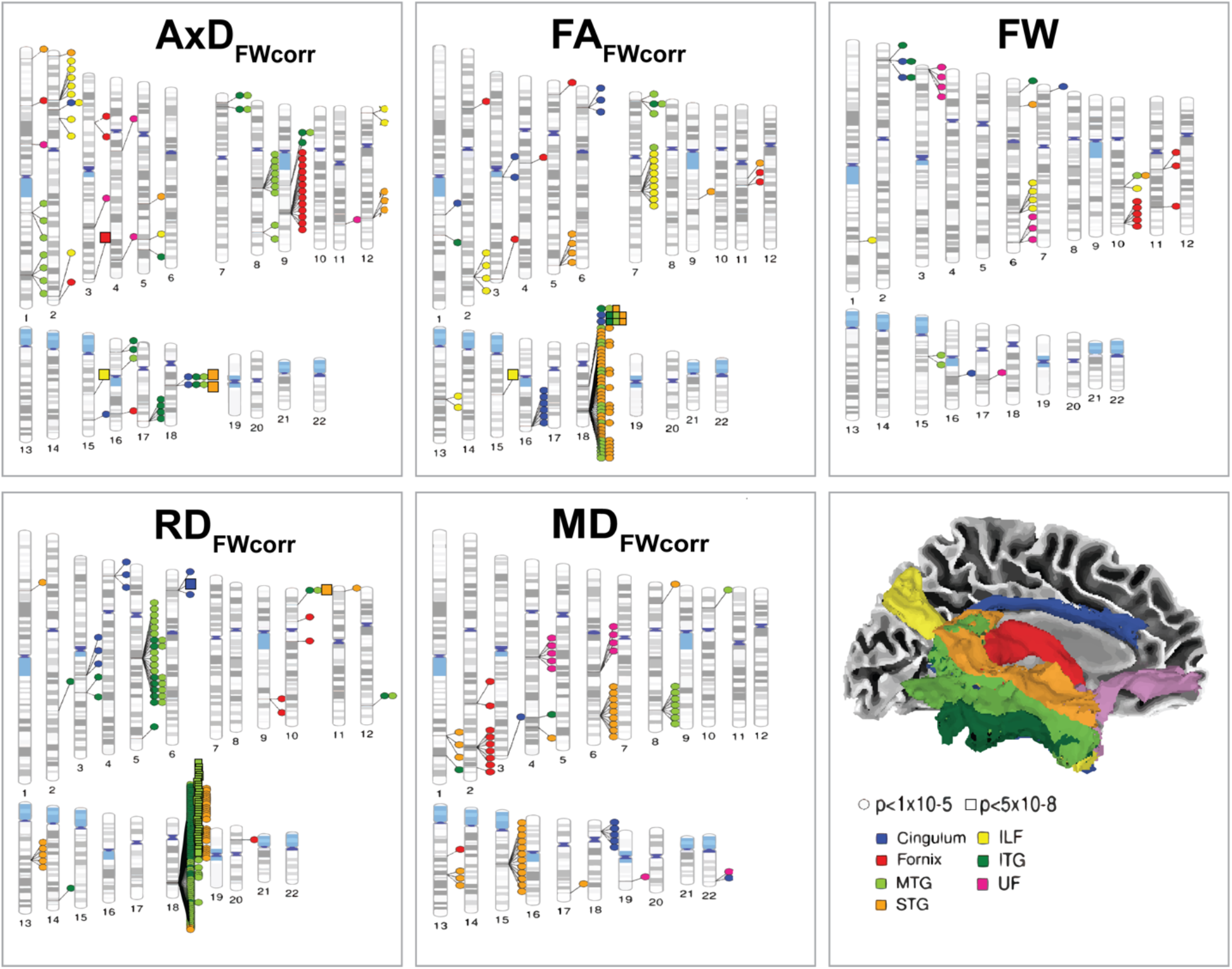
Ideogram of genomic signals associated with WM microstructure. Ideogram of selected genomic regions associated with dMRI metrics. Shape codes for the p-value threshold (*p* < 5×10^−8^; *p* < 1×10^−5^), color codes for the WM tract. The 500 associations between genetic variants and dMRI metrics with the smallest p-value are represented in this plot. This figure was created using PhenoGram from Richie Lab Visualizations (https://visualization.ritchielab.org/phenograms/plot). Abbreviations: AxD, axial diffusivity; FA, fractional anisotropy; FW, free water; ILF, inferior longitudinal fasciculus; ITG, inferior temporal gyrus transcallosal tract; MD, mean diffusivity; MTG, middle temporal gyrus transcallosal tract; RD, radial diffusivity; STG, superior temporal gyrus transcallosal tract; UF, uncinate fasciculus.

**FIGURE 3.**
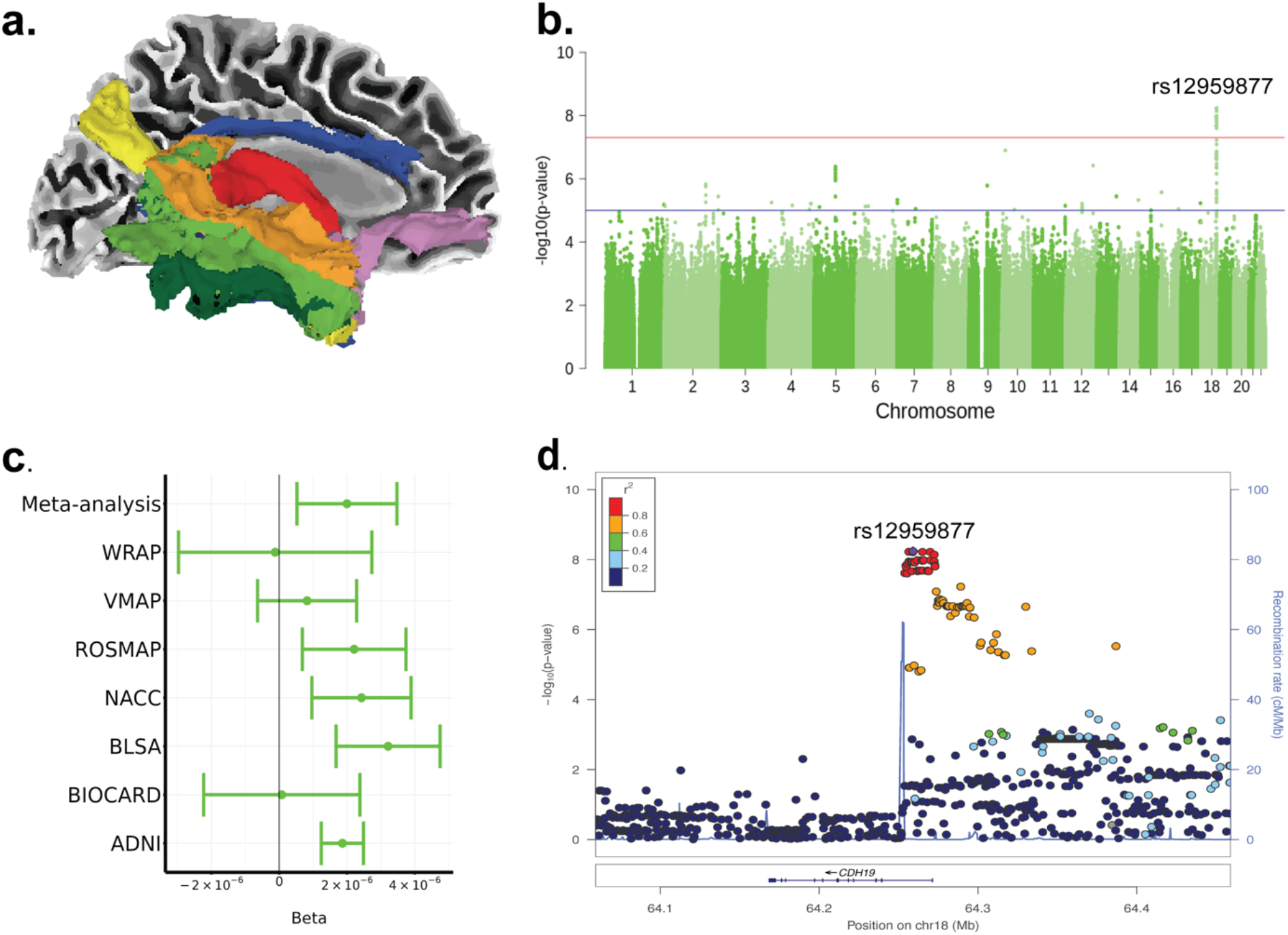
Genome-wide significant locus near *CDH19* associated with MTG RD_Fwcorr_. **a**. Location of the 7 limbic WM in the brain, with the MTG colored in light green. **b**. Manhattan plot of GWAS results for MTG RD_FWcorr_. The red line indicates genome-wide significance (*p* < 5×10^−8^). The blue line indicates suggestive significance (*p* < 1×10^−5^). **c**. Forest plot for the effect of variant rs12959877 for each cohort. **d**. LocusZoom plot for variant rs12959877 on chromosome 18. Abbreviations: ADNI, Alzheimer’s Disease Neuroimaging Initiative; BIOCARD, Predictors of Cognitive Decline Among Normal Individuals; BLSA, Baltimore Longitudinal Study of Aging; chr, chromosome; GWAS, genome-wide association study; MAP, Memory and Aging Project; MTG, middle temporal gyrus transcallosal tract; NACC, National Alzheimer’s Coordinating Center; VMAP, Vanderbilt Memory and Aging Project; ROS, Religious Orders Study; WRAP, Wisconsin Registry for Alzheimer’s Prevention.

We validated the identified variants in the Oxford Brain Imaging Genetics Server (BIG40)^50^ and ENIGMA-VIS.^51^ All of our genome-wide variants were associated with various brain traits such as WM tracts, resting-state functional MRI measures, cortical thickness, and regional and tissue volume (*p* < 0.05, **Table S4c**).

### Significant associations between gene expression of *RORA, FAM107B*, and *KC6* with cognitive function and AD pathology

To evaluate the AD relevance of the genes identified in the GWAS analysis, we investigated the associations of *CDH19, KC6, SENP5, RORA, FAM107B*, and *MIR548A1* expression profiles in the PCC, DLPFC, and CN with cognitive measures (cross-sectional and longitudinal cognition) and AD pathologies (tau tangles, neuritic plaques, amyloid-β, neurofibrillary tangles).

Significant associations were observed for *RORA, FAM107B*, and *KC6* (**Figure 4, Figure S1**). Associations for *RORA* expression included cross-sectional cognition in CN (β = -0.654 ± 0.173; *p*_*FDR*_ = 0.011) and DLPFC (β = -0.570 ± 0.145; *p*_*FDR*_ = 0.001), longitudinal cognition in CN (β = -0.082 ± 0.016; *p*_*FDR*_ = 4.39×10^−5^) and DLPFC (β = -0.570 ± 0.014; *p*_*FDR*_ = 1.31×10^−3^). Significant AD pathologies for *RORA* included tau tangles in CN (β = 0.750944 ± 0.204; *p*_*FDR*_ = 0.010) and DLPFC (β = 0.756 ± 0.165; *p*_*FDR*_ = 7.95×10^−5^), neurite plaques in CN (β = 0.294 ± 0.081; *p*_*FDR*_ = 0.010) and DLPFC (β = 0.165 ± 0.066; *p*_*FDR*_ = 0.049), neurofibrillary tangles in DLPFC (β = 0.194 ± 0.051; *p*_*FDR*_ = 0.002) and amyloid-β in DLPFC (β = 0.515 ± 0.143; *p*_*FDR*_ = 0.003).

**FIGURE 4.**
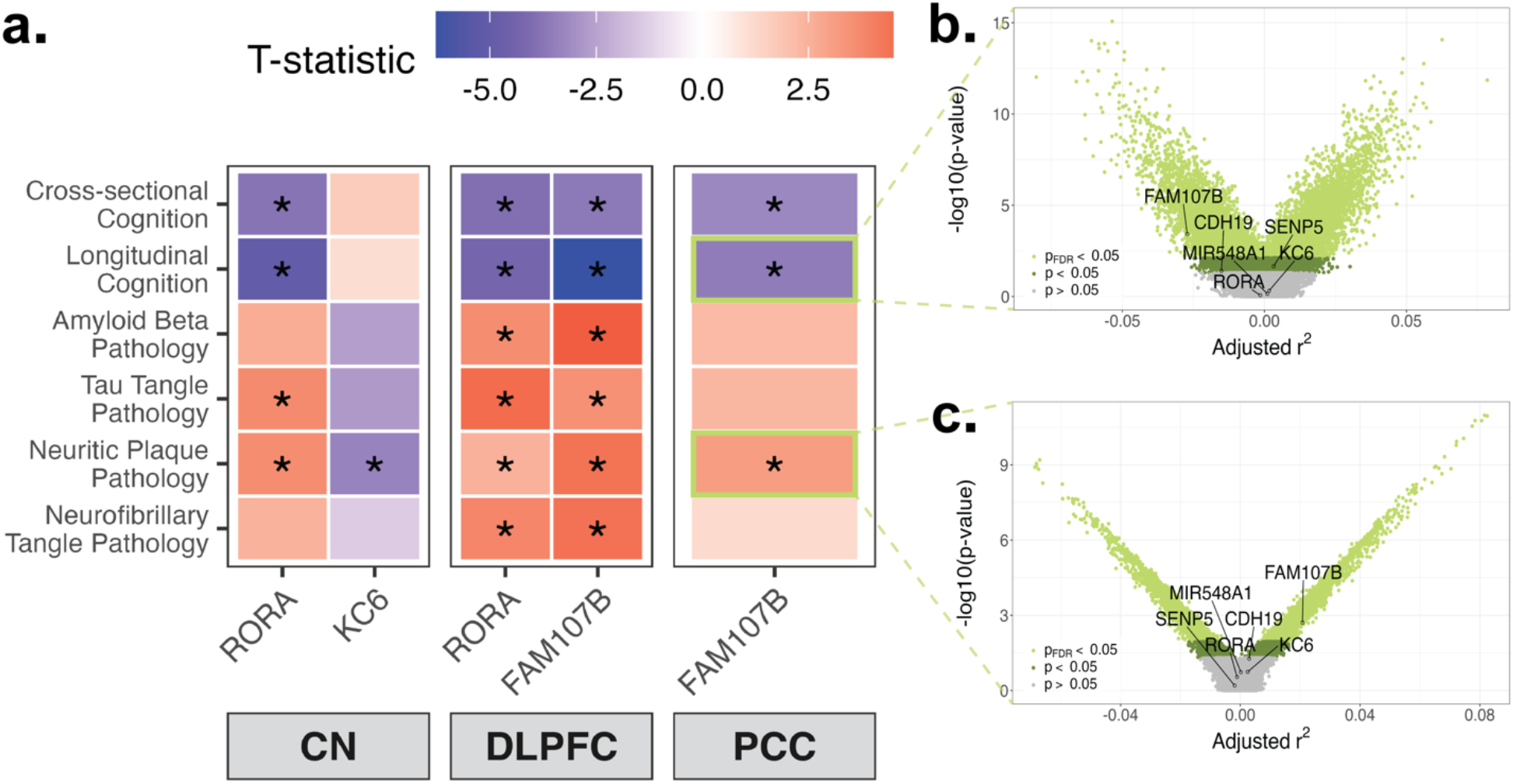
Gene expression profiles in brain tissues of genes identified through GWAS. **a**. Heatmap displaying t-statistics for associations between gene expression profiles in three brain tissues for GWAS-identified genes (*RORA, SENP5, KCNK6, CDH19, FAM107B*, and *MIR548A1*) with cognitive outcomes and AD pathologies. Only genes with at least one significant association (*p*_*FDR*_ < 0.05) with a cognitive or AD outcome are included. **b.–c**. Exemplary volcano plots showing the association of *FAM107B* expression in PCC with **b**. longitudinal cognitive performance and **c**. neuritic plaque burden. Significance thresholds are indicated by color. All volcano plots can be found in **Figure S1**. Abbreviations: AD, Alzheimer’s Disease; CN, caudate nucleus; DLPFC, dorsolateral prefrontal cortex; FDR, false discovery rate; GWAS, genome-wide association study; PCC, posterior cingulate cortex.

For *FAM107B* gene expression, there were significant association with cross-sectional cognition in DLPFC (β = -0.242 ± 0.066; *p*_*FDR*_ = 0.003) and PCC (β = -0.259 ± 0.081; *p*_*FDR*_ = 0.015), longitudinal cognition in DLPFC (β = -0.038 ± 0.006; *p*_*FDR*_ = 2.21×10^−8^) and PCC (β = -0.025 ± 0.007; *p*_*FDR*_ = 0.005). AD pathologies included tau tangles in DLPFC (β = 0.263 ± 0.075; *p*_*FDR*_ = 0.003), neuritic plaques in DLPFC (β = 0.130 ± 0.030; *p*_*FDR*_ = 3.43×10^−4^) and PCC (β = 0.119 ± 0.038; *p*_*FDR*_ = 0.018), neurofibrillary tangles in DLPFC (β = 0.102 ± 0.023; *p*_*FDR*_ = 2.55×10^−4^) and amyloid-β in DLPFC (β = 0.325 ± 0.065; *p*_*FDR*_ = 3.98×10^−5^). Furthermore, *KC6* expression in CN was associated with neurite plaques (β = -0.055 ± 0.017; *p*_*FDR*_ = 0.019). All statistical analyses can be found in **Table S5b**.

### Gene- and pathway-level analyses highlight biological mechanisms related to immune function, neurotrophic signaling, and cardiovascular traits

The genetic architecture of WM microstructure was also investigated at the gene and pathway level. Gene-level analyses revealed a significant association between cingulum AxD_FWcorr_ and the gene *SERPINA12* (z = 4.60, *p*_*FDR*_ = 0.038, N_SNPs_ = 132) which is known to influence obesity and atherosclerosis by modulating glycolipid metabolism.^59–61^ **Table S6** depicts the statistics for all gene analyses.

Pathway analysis identified significant pathways (*p*_*FDR*_ < 0.05) for ITG FA_FWcorr_, STG FA_FWcorr_, and UF AxD_FWcorr_. Enriched pathways for ITG FA_FWcorr_ were related to immune function, encompassing isotype switching of B cells, B cell-mediated immunity, and the production of immunoglobulins and antibodies. Beyond immune-related pathways, STG FA_FWcorr_ was associated with several neurotrophic signaling pathways involving NTRK3 and NTRK2 via the RAS pathway, which are crucial for nervous system development and survival. This tract was also linked to insulin-like growth factor 1 receptor (IGF1R) signaling and insulin receptor binding pathways. Furthermore, STG FA_FWcorr_ showed associations with cancer mechanisms, including mutant forms and internalization of the epidermal growth factor receptor (EGFR). It also included pathways related to the overexpression of the human epidermal growth factor receptor 2 (HER2) and signaling events mediated by the EphA2 receptor. For UF AxD_FWcorr_, enriched pathways included photoreceptor cell outer segment organization and the binding of the peptide hormone angiotensin to its receptor. Statistics for all pathway analysis can be found in **Table S7**.

### Genetic covariance between WM microstructure with cardiovascular, lipid, inflammatory, and neurological traits

To investigate the extent of shared genetic factors between WM microstructure and complex traits (N = 65), we performed genetic covariance analyses using GNOVA. Traits that exhibited at least one FDR-corrected significant association with one dMRI metric are displayed in **Figure 5a**. HDL cholesterol demonstrated multiple significant associations with WM microstructure. Specifically, we observed negative covariance with FA_FWcorr_ and AxD_FWcorr_, alongside positive covariance with MD_FWcorr_, RD_FWcorr_, and FW. When examining the local genetic covariance for HDL cholesterol with MD_FWcorr_ and RD_FWcorr_, multiple distributed genomic regions exhibiting significant covariation were identified (**Figure 5b, Table S8**).

**FIGURE 5.**
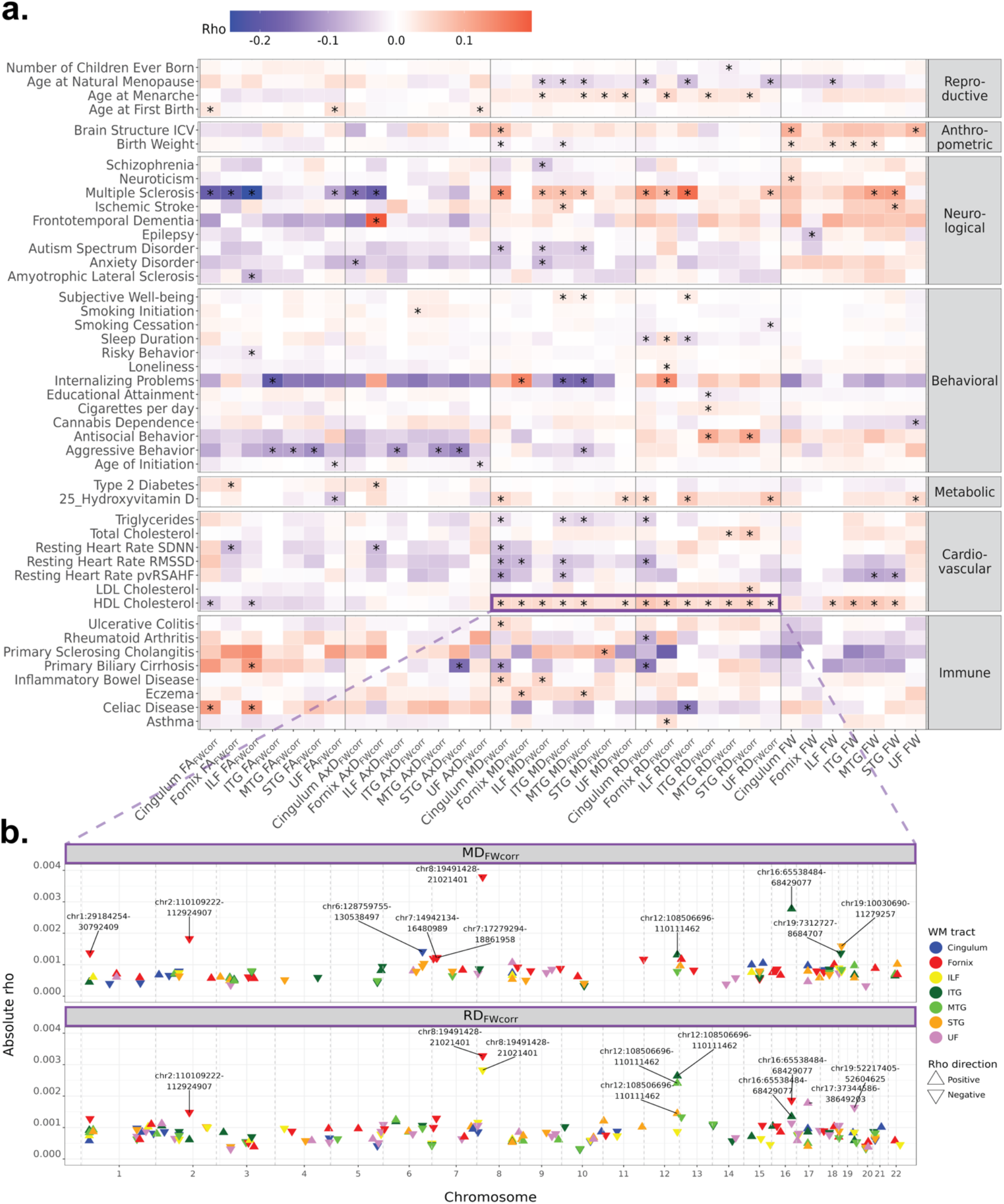
Genetic covariance between complex traits and WM microstructure. **a**. Genetic covariance between dMRI metrics and complex traits which have shown at least one association with a dMRI metric (*p*_*FDR*_ < 0.05). “*” marks a *p*_*FDR*_ < 0.05. **b**. Local genetic covariance between HDL cholesterol with MD_FWcorr_ and RD_FWcorr_. Only genomic regions with a *p* < 0.05 were included. Abbreviations: AxD, axial diffusivity; FA, fractional anisotropy; FDR, false discovery rate; FW, free water; ILF, inferior longitudinal fasciculus; ITG, inferior temporal gyrus transcallosal tract; MD, mean diffusivity; MTG, middle temporal gyrus transcallosal tract; RD, radial diffusivity; STG, superior temporal gyrus transcallosal tract; UF, uncinate fasciculus.

Resting heart rate variability traits (SDNN, RMSSD, pvRSAHF) and triglyceride showed negative associations with MD_FWcorr_, RD_FWcorr_, and FW. For metabolic traits, type 2 diabetes was positively associated with fornix FA_FWcorr_ and AxD_FWcorr_, whereas hydroxyvitamin D demonstrated several positive associations with MD_FWcorr_ and RD_FWcorr_. Moreover, multiple genetic associations between WM microstructure were identified with immune-related diseases, including ulcerative colitis, rheumatoid arthritis, primary sclerosing cholangitis, primary biliary cirrhosis, inflammatory bowel disease, eczema, celiac disease, and asthma. Neurological and psychiatric traits also showed significant associations, encompassing schizophrenia, neuroticism, ischemic stroke, frontotemporal dementia, epilepsy, autism spectrum disorder, anxiety disorder, and amyotrophic lateral sclerosis. Behavioral traits such as subjective well-being, smoking initiation, smoking cessation, cigarettes per day, cannabis dependence, sleep duration, risky behavior, loneliness, internalizing problems, educational attainment, antisocial behavior, aggressive behavior, and age of initiation were linked to WM microstructure. Statistics for all computed tests can be found in **Table S9b**. Most of the genetic covariance results remained significant and showed similar effect directions after removing the *APOE* region from the genome (**Figure S2, Table S10**).

Genetic covariance analysis between FW-corrected dMRI metrics and 3,143 neuroimaging-derived phenotypes from the UK Biobank identified 9,218 significant associations after FDR correction out of 110,005 tests performed (**Table S11**). **Figure S3** highlights the neuroimaging-derived phenotypes with at least 10 significant associations across the FW-corrected dMRI metrics. Notably, substantial genetic overlap was observed with the UK Biobank dMRI measures (e.g., mode of the arcuate fasciculus, FA_CONV_ of the posterior corona radiata) and resting-state network metrics (e.g., NET100_0452, NET100_1425).

### TWAS identified an association between STG RD_FWcorr_ and *DNAJB14*

The summary statistics derived from the meta-analyzed GWAS of WM microstructure were used to compute TWAS using S-PrediXcan. A significant association between STG RD_FWcorr_ and the gene *DNAJB14* in colon transverse tissue (z = -5.47, *p*_*FDR*_ = 0.016, N_SNPs_ = 27) was identified. *DNAJB14* is involved in chaperone cofactor-dependent protein refolding and protein-containing complex assembly.^62^ Results (*p* < 0.05) for all analyses can be found in **Table S12**.

## Discussion

This study investigated the genetic architecture of limbic WM microstructure in a large, harmonized sample of older adults enriched for cognitive impairment, utilizing advanced FW-corrected dMRI metrics. We observed substantial heritability of limbic WM microstructure, especially within tracts such as the cingulum, fornix and ILF. Our meta-analyzed GWAS identified several genetic loci associated with WM characteristics, implicating the genes *CDH19, KC6, SENP5, RORA, FAM107B*, and *MIR548A1*. Furthermore, we identified gene-level association of WM microstructure with *DNAJB14* using TWAS and with *SERPINA12* using gene-level analysis. Pathway and genetic covariance analyses revealed significant links between limbic WM microstructure and biological processes related to inflammation, vascular health, lipid metabolism, and various neurological conditions.

The strongest GWAS signal identified was a locus including 38 SNPs on chromosome 18, with most SNPs acting as eQTLs for *CDH19*. This gene, also known as Cadherin 19, encodes a calcium-dependent cell-adhesion glycoprotein highly expressed in oligodendrocytes and Schwann cells, which are crucial for myelin formation and maintenance.^58^ This finding suggests a link between genetic variation influencing myelin-related cell adhesion processes and WM microstructural integrity in aging. Another notable association was found near *SENP5* on chromosome 3 with fornix AxD_FWcorr_. *SENP5* regulates SUMOylation, a post-translational modification critical for gene expression, DNA repair, and mitochondrial dynamics, and has been implicated in synaptogenesis and synaptic function in mature neurons.^63,64^

Several other GWAS-identified genes, including *RORA, FAM107B* and *KC6*, showed significant association with their brain tissue expression levels, cognitive decline and AD pathologies (tau tangles, neurite plaques, neurofibrillary tangles and amyloid-β plaques) in our bulk RNA-seq analyses. *RORA* encodes a protein that binds to hormone response elements upstream of multiple genes to enhance their expression. It regulates key genes involved in neurological functions and is implicated in autism spectrum disorders, highlighting its role in neuronal differentiation and synaptic plasticity.^65^ As a nuclear receptor transcription factor, *RORA* interacts with transcriptional regulators like insulin and brain-derived neurotrophic factors, both associated with AD.^66^ Additionally, *RORA* supports neuronal survival and has a neuroprotective role in Parkinson’s Disease by protecting neurons from oxidative stress.^67^ *FAM107B* is widely expressed in the brain and associated with various brain structural features, including cortical thickness, subcortical volume, and overall brain morphology.^68^ *KC6* is a less well-characterized RNA gene and has been associated with corneal biology and diseases.^69^ This evidence indicates that some of the identified genes are involved in general brain health, which the observed genetic associations between WM microstructure and neurological and psychiatric conditions such as multiple sclerosis, aggressive behavior, and internalizing problems further supported.

Our study strongly implicates inflammatory, lipid metabolism, and vascular mechanisms in limbic WM health. *RORA* is also involved in regulating cholesterol and glucose metabolism and blood vessel morphogenesis.^66^ *SENP5*, in addition to its importance for brain health, has a critical role in cardiac function by regulating mitochondrial balance. Excessive de-SUMOylation by *SENP5*, which is upregulated in human heart failure, has been linked to cardiomyopathies and cardiac dysfunction.^70,71^ Our post-GWAS analyses supported associations between WM microstructure with lipid and vascular mechanisms, revealing gene-level associations for *SERPINA12* and links to insulin and inflammation pathways. Notably, *SERPINA12* is elevated in individuals with obesity and is associated with obesity-linked metabolic traits, such as insulin resistance, impaired glycemic control, and cardiovascular disease.^72^ The connection between WM with lipid and vascular mechanisms was also demonstrated by genetic overlap between WM microstructure and HDL cholesterol, triglycerides, resting heart rate variability traits, ischemic stroke, type 2 diabetes, and several inflammatory diseases. These findings suggest that cardiovascular risk and associated systemic inflammation may provide a pathophysiological basis for WM alterations observed in later life, which can in turn contribute to risk for neurodegenerative diseases and cognitive decline. Vascular conditions such as hypertension, atherosclerosis, and type 2 diabetes may compromise vascular integrity, leading to reduced blood flow or microvascular damage in the brain. Such vascular challenges adversely affect WM, potentially resulting in axonal and myelin damage. This highlights the importance of intact energy metabolism and the cardiovascular system in brain health. Further research is needed to disentangle signals related to vascular mechanisms from AD specific pathologies.

A key strength of this study is the investigation of genetic influence on WM microstructure within well-characterized cohorts of older individuals, including those with cognitive impairment, which enhances our ability to uncover biological mechanisms relevant to aging and in neurodegenerative diseases. Unlike large-scale studies such as UK Biobank and ENIGMA that have predominantly focused on WM in midlife, our study specifically addresses the genetic architecture of WM degeneration in the context of aging and AD risk. Additionally, the use of FW-corrected dMRI metrics provides a more accurate quantification of WM microstructure by mitigating confounds from extracellular free water.

This study has several limitations. Our analyses were conducted on non-Hispanic White individuals, which, while reducing confounding from population stratification, limits the generalizability of our findings to more diverse populations. Future research should aim to replicate these findings in larger, multi-ethnic cohorts. The cross-sectional and correlational nature of our analyses precludes causal inferences. While the use of single-shell dMRI data allowed for the inclusion of a large, harmonized dataset, future studies employing multi-shell dMRI approaches, such as NODDI,^73^ could provide more detailed insights into distinct tissue compartments and potentially quantify aspects like neuroinflammation. Finally, only a small subset of our cohort (5%) had a formal clinical diagnosis of AD at the time of imaging. Enriching future cohorts with individuals across the AD spectrum may enhance the detection of neurodegeneration-specific genetic signals.

## Conclusion

This imaging genetics study identified several novel genes linked to limbic WM microstructure in multiple, harmonized aging cohorts. Notably, bulk RNA-seq analyses demonstrated that some of these genes were associated with cognitive decline and amyloid-β/tau burden, suggesting AD relevance. Furthermore, our findings revealed that the genetic architecture of limbic WM is strongly tied to vascular health and inflammation, highlighting these pathways as promising avenues for future therapeutic development

## Data availability

Data from ADNI, NACC, ROSMAP, and WRAP is available on NIAGADS (https://dss.niagads.org/). For BIOCARD (https://biocard.pathology.jhu.edu/resources-for-researchers/), BLSA (https://www.blsa.nih.gov/), and VMAP (https://vmacdata.org/vmap), data use can be approved through each cohort’s website. The WM GWAS summary statistics generated in this study will be made available on NIAGADS.

## Funding Acknowledgments

This study was supported by several funding sources, including K01-EB032898 (KGS), R01-EB017230 (BAL) K01-AG073584 (DBA), U24-AG074855 (TJH), 75N95D22P00141 (TJH), R01-AG059716 (TJH), UL1-TR000445 and UL1-TR002243 (Vanderbilt Clinical Translational Science Award), S10-OD023680 (Vanderbilt’s High-Performance Computer Cluster for Biomedical Research). The research was support in part by the Intramural Research Program of the National Institutes of Health, National Institute on Aging. Study data were obtained from the Vanderbilt Memory and Aging Project (VMAP). VMAP data were collected by Vanderbilt Memory and Alzheimer’s Center Investigators at Vanderbilt University Medical Center. This work was supported by NIA grants R01-AG034962 (PI: Jefferson), R01-AG056534 (PI: Jefferson), U19-AG03655 (PI:Albert) and Alzheimer’s Association IIRG-08-88733 (PI: Jefferson). The data contributed from the Wisconsin Registry for Alzheimer’s Prevention was supported by NIA AG021155, AG027161, AG037639, and AG054047. The BLSA is supported by the Intramural Research Program of the National Institutes of Health, National Institute on Aging. This research was supported in part by the Intramural Research Program of the National Institutes of Health, National Institute on Aging. Data collection and sharing for this project was funded (in part) by the Alzheimer’s Disease Neuroimaging Initiative (ADNI) (National Institutes of Health Grant U01 AG024904) and DOD ADNI (Department of Defense award number W81XWH-12-2-0012). ADNI is funded by the National Institute on Aging, the National Institute of Biomedical Imaging and Bioengineering, and through generous contributions from the following: AbbVie, Alzheimer’s Association; Alzheimer’s Drug Discovery Foundation; Araclon Biotech; BioClinica, Inc.; Biogen; Bristol-Myers Squibb Company; CereSpir, Inc.; Cogstate; Eisai Inc.; Elan Pharmaceuticals, Inc.; Eli Lilly and Company; EuroImmun; F. Hoffmann-La Roche Ltd and its affiliated company Genentech, Inc.; Fujirebio; GE Healthcare; IXICO Ltd.; Janssen Alzheimer Immunotherapy Research & Development, LLC.; Johnson & Johnson Pharmaceutical Research & Development LLC.; Lumosity; Lundbeck; Merck & Co., Inc.; Meso Scale Diagnostics, LLC.; NeuroRx Research; Neurotrack Technologies; Novartis Pharmaceuticals Corporation; Pfizer Inc.; Piramal Imaging; Servier; Takeda Pharmaceutical Company; and Transition Therapeutics. The Canadian Institutes of Health Research is providing funds to support ADNI clinical sites in Canada. Private sector contributions are facilitated by the Foundation for the National Institutes of Health (www.fnih.org). The grantee organization is the Northern California Institute for Research and Education, and the study is coordinated by the Alzheimer’s Therapeutic Research Institute at the University of Southern California. ADNI data are disseminated by the Laboratory for Neuro Imaging at the University of Southern California. Data contributed from MAP/ROS was supported by NIA R01AG017917, P30AG10161, P30AG072975, R01AG056405, UH2NS100599, UH3NS100599, R01AG064233, R01AG015819 and R01AG067482, and the Illinois Department of Public Health (Alzheimer’s Disease Research Fund). Data can be accessed at www.radc.rush.edu. The NACC database is funded by NIA/NIH Grant U24 AG072122. NACC data are contributed by the NIA-funded ADCs : P50 AG005131 (PI James Brewer, MD, PhD), P50 AG005133 (PI Oscar Lopez, MD), P50 AG005134 (PI Bradley Hyman, MD, PhD), P50 AG005136 (PI Thomas Grabowski, MD), P50 AG005138 (PI Mary Sano, PhD), P50 AG005142 (PI Helena Chui, MD), P50 AG005146 (PI Marilyn Albert, PhD), P50 AG005681 (PI John Morris, MD), P30 AG008017 (PI Jeffrey Kaye, MD), P30 AG008051 (PI Thomas Wisniewski, MD), P50 AG008702 (PI Scott Small, MD), P30 AG010124 (PI John Trojanowski, MD, PhD), P30 AG010129 (PI Charles DeCarli, MD), P30 AG010133 (PI Andrew Saykin, PsyD), P30 AG072975 (PI Julie Schneider, MD), P30 AG012300 (PI Roger Rosenberg, MD), P30 AG013846 (PI Neil Kowall, MD), P30 AG013854 (PI Robert Vassar, PhD), P50 AG016573 (PI Frank LaFerla, PhD), P50 AG016574 (PI Ronald Petersen, MD, PhD), P30 AG019610 (PI Eric Reiman, MD), P50 AG023501 (PI Bruce Miller, MD), P50 AG025688 (PI Allan Levey, MD, PhD), P30-AG072946 (PI Linda Van Eldik, PhD), P50 AG033514 (PI Sanjay Asthana, MD, FRCP), P30 AG035982 (PI Russell Swerdlow, MD), P50 AG047266 (PI Todd Golde, MD, PhD), P50 AG047270 (PI Stephen Strittmatter, MD, PhD), P50 AG047366 (PI Victor Henderson, MD, MS), P30 AG049638 (PI Suzanne Craft, PhD), P30 AG053760 (PI Henry Paulson, MD, PhD), P30 AG066546 (PI Sudha Seshadri, MD), P20 AG068024 (PI Erik Roberson, MD, PhD), P20 AG068053 (PI Marwan Sabbagh, MD), P20 AG068077 (PI Gary Rosenberg, MD), P20 AG068082 (PI Angela Jefferson, PhD), P30 AG072958 (PI Heather Whitson, MD), P30 AG072959 (PI James Leverenz, MD). NACC data can be accessed at naccdata.org.

## Consent Statement

All participants provided informed consent in their respective cohort studies.

## SUPPLEMENTARY FIGURES

**FIGURE S1.**
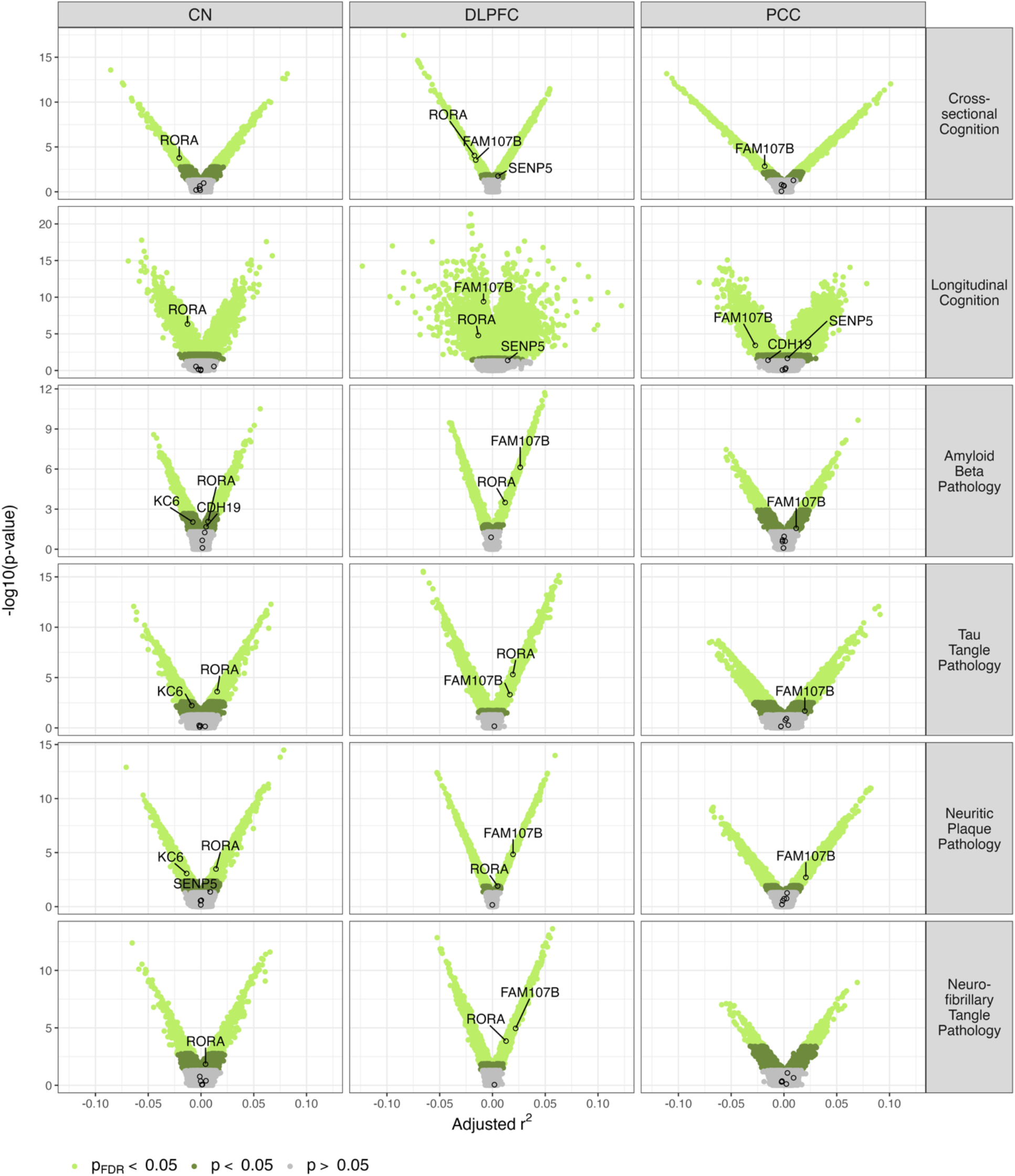
Gene expression profiles in brain tissues of genes identified through GWAS. Volcano plots of gene expression profiles in brain tissues for the genes *RORA, SENP5, KC6, CDH19, FAM107B*, and *MIR548A1* identified through GWAS associated with cognitive outcomes and AD pathologies. Significance thresholds are indicated by color. Highlighted genes with a *p* < 0.05 are labeled. Abbreviations: AD, Alzheimer’s Disease; CN, caudate nucleus; DLPFC, dorsolateral prefrontal cortex; GWAS, genome-wide association study; FDR, false discovery rate; PCC, posterior cingulate cortex.

**FIGURE S2.**
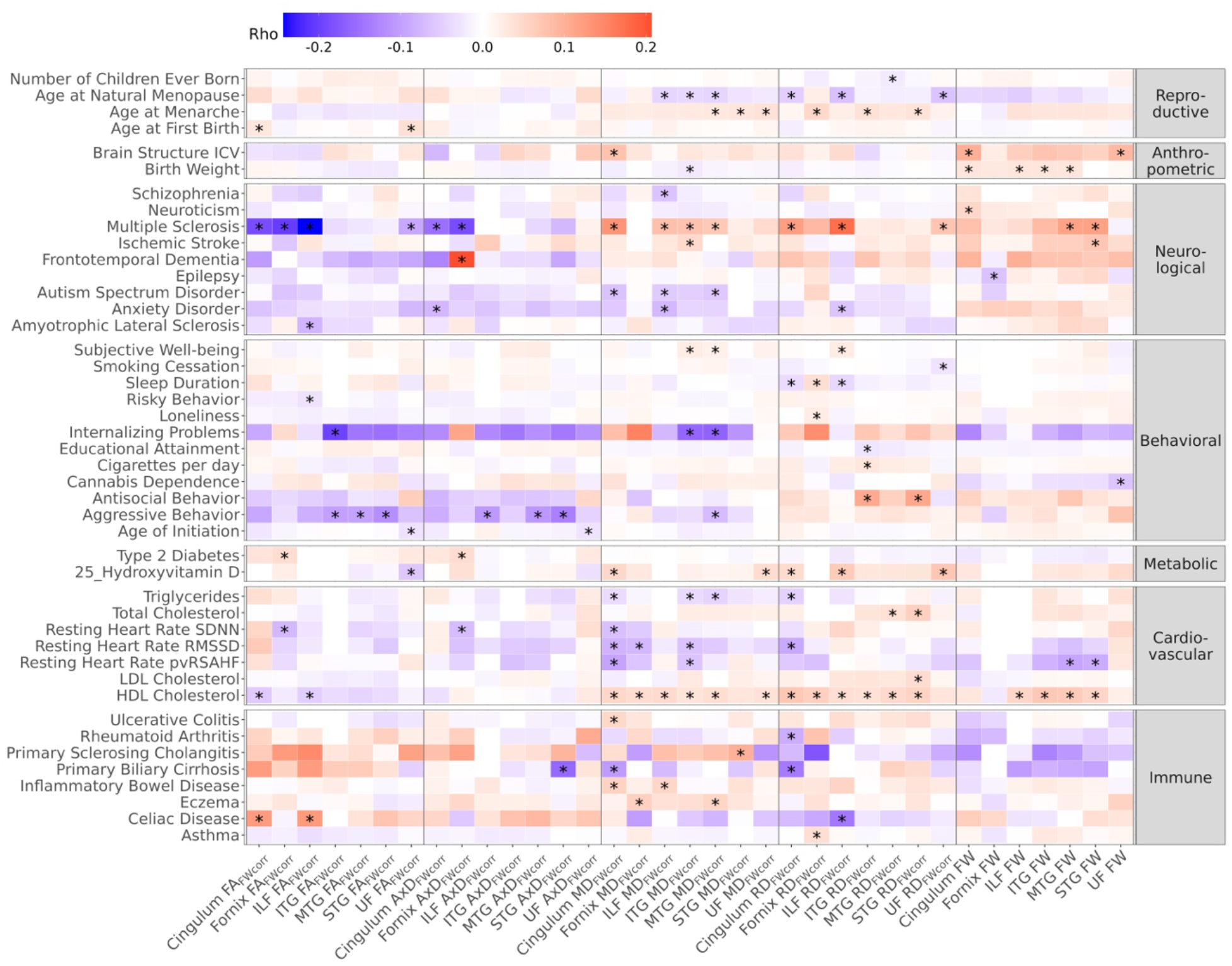
Genetic covariance between complex traits and WM microstructure when the *APOE* region is removed from the genome. Genetic covariance between dMRI metrics (x-axis) and complex traits and diseases (y-axis) when the *APOE* region is removed from the genome. The traits included have shown at least one FDR-significant association with a dMRI metric. “*” marks genetic covariance with a *p*_*FDR*_ < 0.05. Abbreviations: AxD, axial diffusivity; FA, fractional anisotropy; FDR, false discovery rate; FW, free water; ILF, inferior longitudinal fasciculus; ITG, inferior temporal gyrus transcallosal tract; MD, mean diffusivity; MTG, middle temporal gyrus transcallosal tract; RD, radial diffusivity; STG, superior temporal gyrus transcallosal tract; UF, uncinate fasciculus.

**FIGURE S3.**
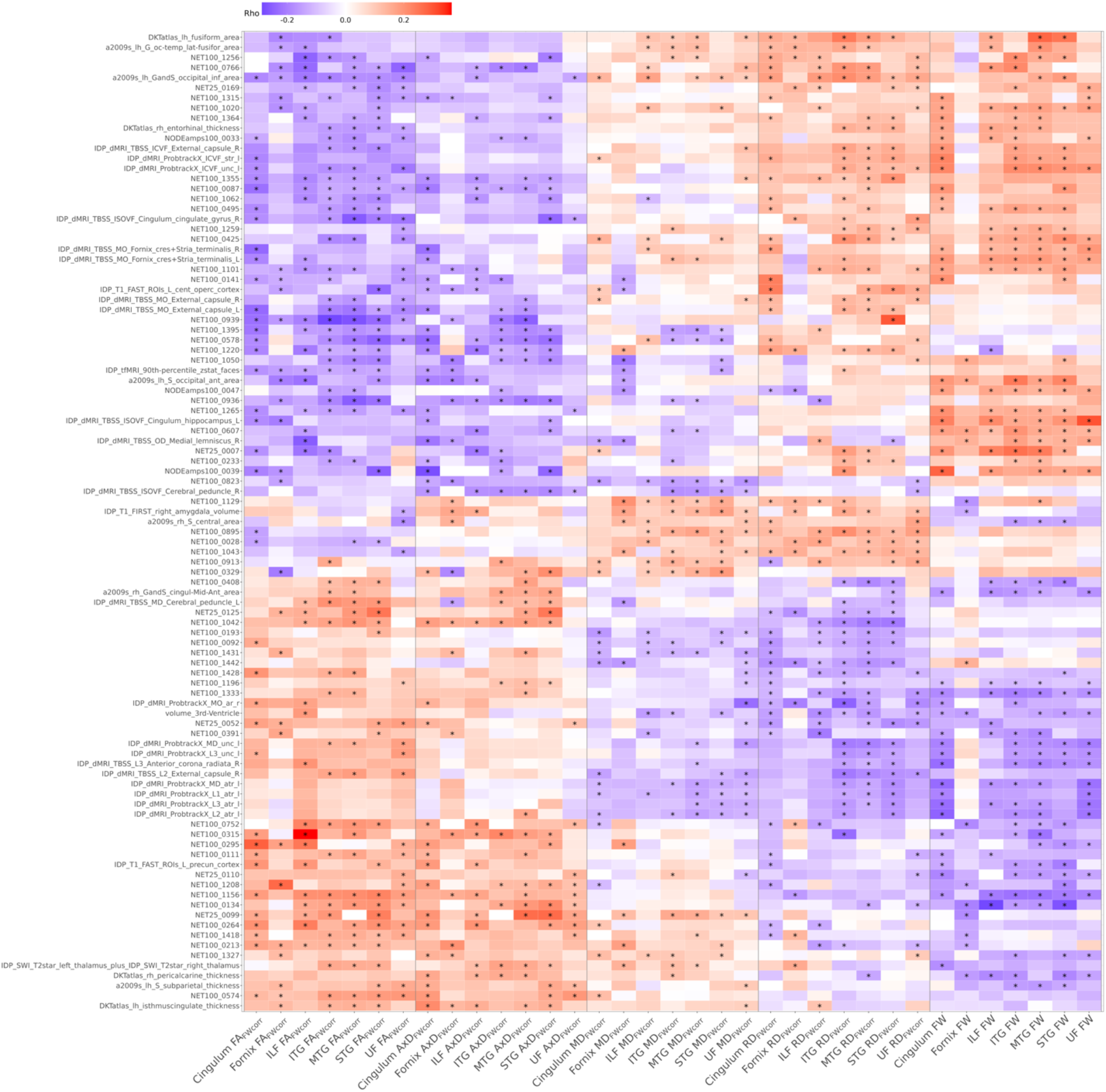
Genetic covariance between brain traits and WM microstructure. Genetic covariance between dMRI metrics (x-axis) and brain traits (y-axis). The traits included have shown at least ten FDR-significant association with a dMRI metric. “*” marks genetic covariance with a *p*_*FDR*_ < 0.05. The y-axis is clustered using Euclidian distance. Abbreviations: AxD, axial diffusivity; FA, fractional anisotropy; FDR, false discovery rate; FW, free water; ILF, inferior longitudinal fasciculus; ITG, inferior temporal gyrus transcallosal tract; MD, mean diffusivity; MTG, middle temporal gyrus transcallosal tract; RD, radial diffusivity; STG, superior temporal gyrus transcallosal tract; UF, uncinate fasciculus.

## References

1. Penke, L. et al. Brain-wide white matter tract integrity is associated with information processing speed and general intelligence. Mol Psychiatry 17, 955–955 (2012).

2. Ritchie, S. J. et al. Coupled Changes in Brain White Matter Microstructure and Fluid Intelligence in Later Life. J Neurosci 35, 8672–8682 (2015).

3. Cox, S. R. et al. Ageing and brain white matter structure in 3,513 UK Biobank participants. Nat Commun 7, 13629 (2016).

4. Ihara, M. et al. Quantification of myelin loss in frontal lobe white matter in vascular dementia, Alzheimer’s disease, and dementia with Lewy bodies. Acta Neuropathol 119, 579–589 (2010).

5. Sachdev, P. S., Zhuang, L., Braidy, N. & Wen, W. Is Alzheimer’s a disease of the white matter? Current Opinion in Psychiatry 26, 244 (2013).

6. Simpson, J. E. et al. Microglial activation in white matter lesions and nonlesional white matter of ageing brains. Neuropathology and Applied Neurobiology 33, 670–683 (2007).

7. Yang, Y. et al. White matter microstructural metrics are sensitively associated with clinical staging in Alzheimer’s disease. Alzheimer’s & Dementia: Diagnosis, Assessment & Disease Monitoring 15, e12425 (2023).

8. Archer, D. B. et al. Leveraging longitudinal diffusion MRI data to quantify differences in white matter microstructural decline in normal and abnormal aging. Alzheimer’s & Dementia: Diagnosis, Assessment & Disease Monitoring 15, e12468 (2023).

9. Peterson, A. et al. Sex and APOE ε4 allele differences in longitudinal white matter microstructure in multiple cohorts of aging and Alzheimer’s disease. Alzheimers Dement (2024) doi:10.1002/alz.14343.

10. Lee, S. et al. White matter hyperintensities are a core feature of Alzheimer’s disease: Evidence from the Dominantly Inherited Alzheimer Network. Ann Neurol 79, 929–939 (2016).

11. Nasrabady, S. E., Rizvi, B., Goldman, J. E. & Brickman, A. M. White matter changes in Alzheimer’s disease: a focus on myelin and oligodendrocytes. Acta Neuropathologica Communications 6, 22 (2018).

12. Archer, D. B. et al. Free-water metrics in medial temporal lobe white matter tract projections relate to longitudinal cognitive decline. Neurobiol Aging 94, 15–23 (2020).

13. Basser, P. J. & Jones, D. K. Diffusion-tensor MRI: theory, experimental design and data analysis - a technical review. NMR Biomed 15, 456–467 (2002).

14. Jenkinson, M., Beckmann, C. F., Behrens, T. E. J., Woolrich, M. W. & Smith, S. M. FSL. Neuroimage 62, 782–790 (2012).

15. Le Bihan, D. et al. MR imaging of intravoxel incoherent motions: application to diffusion and perfusion in neurologic disorders. Radiology 161, 401–407 (1986).

16. Le Bihan, D. & Breton, E. Imagerie de diffusion in-vivo par résonance magnétique nucléaire. Comptes-Rendus de l’Académie des Sciences 93, 27–34 (1985).

17. Pasternak, O., Sochen, N., Gur, Y., Intrator, N. & Assaf, Y. Free water elimination and mapping from diffusion MRI. Magnetic Resonance in Medicine 62, 717–730 (2009).

18. Gustavson, D. E. et al. Exploring the genetics of rhythmic perception and musical engagement in the Vanderbilt Online Musicality Study. Annals of the New York Academy of Sciences 1521, 140–154 (2023).

19. Vuoksimaa, E. et al. Heritability of white matter microstructure in late middle age: A twin study of tract-based fractional anisotropy and absolute diffusivity indices. Hum Brain Mapp 38, 2026–2036 (2016).

20. Zhao, B. et al. Common genetic variation influencing human white matter microstructure. Science 372, (2021).

21. Zhao, B. et al. Large-scale GWAS reveals genetic architecture of brain white matter microstructure and genetic overlap with cognitive and mental health traits (n = 17,706). Mol Psychiatry 26, 3943–3955 (2021).

22. Bagepally, B. S. et al. Apolipoprotein E4 and Brain White Matter Integrity in Alzheimer’s Disease: Tract-Based Spatial Statistics Study under 3-Tesla MRI. Neurodegenerative Diseases 10, 145–148 (2012).

23. De Rossi, P. et al. Predominant expression of Alzheimer’s disease-associated BIN1 in mature oligodendrocytes and localization to white matter tracts. Mol Neurodegener 11, 59 (2016).

24. Harrison, J. R. et al. Imaging Alzheimer’s genetic risk using diffusion MRI: A systematic review. Neuroimage Clin 27, 102359 (2020).

25. Heise, V., Filippini, N., Ebmeier, K. P. & Mackay, C. E. The APOE ?4 allele modulates brain white matter integrity in healthy adults. Mol Psychiatry 16, 908–916 (2011).

26. Foley, S. F. et al. Multimodal Brain Imaging Reveals Structural Differences in Alzheimer’s Disease Polygenic Risk Carriers: A Study in Healthy Young Adults. Biol Psychiatry 81, 154–161 (2017).

27. Lorenz, A. et al. The effect of Alzheimer’s disease genetic factors on limbic white matter microstructure. Alzheimer’s & Dementia 21, e70130 (2025).

28. Jack, C. R. et al. The Alzheimer’s Disease Neuroimaging Initiative (ADNI): MRI methods. Journal of Magnetic Resonance Imaging 27, 685–691 (2008).

29. Albert, M. et al. Cognitive changes preceding clinical symptom onset of mild cognitive impairment and relationship to ApoE genotype. Curr Alzheimer Res 11, 773–784 (2014).

30. Besser, L. M. et al. The Revised National Alzheimer’s Coordinating Center’s Neuropathology Form—Available Data and New Analyses. Journal of Neuropathology & Experimental Neurology 77, 717–726 (2018).

31. Weintraub, S. et al. The Alzheimer’s Disease Centers’ Uniform Data Set (UDS): The Neuropsychologic Test Battery. Alzheimer Disease & Associated Disorders 23, 91 (2009).

32. Weintraub, S. et al. Version 3 of the Alzheimer Disease Centers’ Neuropsychological Test Battery in the Uniform Data Set (UDS). Alzheimer Disease & Associated Disorders 32, 10 (2018).

33. Bennett, D. A. et al. Religious Orders Study and Rush Memory and Aging Project. J Alzheimers Dis 64, S161–S189 (2018).

34. Moore, E. E. et al. Increased Left Ventricular Mass Index Is Associated With Compromised White Matter Microstructure Among Older Adults. Journal of the American Heart Association: Cardiovascular and Cerebrovascular Disease 7, e009041 (2018).

35. Johnson, S. C. et al. The Wisconsin Registry for Alzheimer’s Prevention: A review of findings and current directions. Alzheimers Dement (Amst) 10, 130–142 (2017).

36. Sager, M. A., Hermann, B. & La Rue, A. Middle-Aged Children of Persons With Alzheimer’s Disease: APOE Genotypes and Cognitive Function in the Wisconsin Registry for Alzheimer’s Prevention. J Geriatr Psychiatry Neurol 18, 245–249 (2005).

37. Cai, L. Y. et al. PreQual: An automated pipeline for integrated preprocessing and quality assurance of diffusion weighted MRI images. Magn Reson Med 86, 456–470 (2021).

38. Schilling, K. G. et al. Synthesized b0 for diffusion distortion correction (Synb0-DisCo). Magn Reson Imaging 64, 62–70 (2019).

39. Avants, B. B., Epstein, C. L., Grossman, M. & Gee, J. C. Symmetric diffeomorphic image registration with cross-correlation: evaluating automated labeling of elderly and neurodegenerative brain. Med Image Anal 12, 26–41 (2008).

40. Kim, M. E. et al. Scalable, reproducible, and cost-effective processing of large-scale medical imaging datasets. in Medical Imaging 2025: Imaging Informatics vol. 13411 128–140 (SPIE, 2025).

41. Archer, D. B., Coombes, S. A., McFarland, N. R., DeKosky, S. T. & Vaillancourt, D. E. Development of a transcallosal tractography template and its application to dementia. Neuroimage 200, 302–312 (2019).

42. Brown, C. A. et al. Development, validation and application of a new fornix template for studies of aging and preclinical Alzheimer’s disease. Neuroimage Clin 13, 106–115 (2017).

43. Archer, D. Tractography Templates for White Matter Microstructure Analysis in Aging and Alzheimer’s Disease. Zenodo 10.5281/zenodo.13863955 (2024).

44. Beer, J. C. et al. Longitudinal ComBat: A method for harmonizing longitudinal multi-scanner imaging data. Neuroimage 220, 117129 (2020).

45. Eissman, J. M. et al. Sex differences in the genetic architecture of cognitive resilience to Alzheimer’s disease. Brain 145, 2541–2554 (2022).

46. Taliun, D. et al. Sequencing of 53,831 diverse genomes from the NHLBI TOPMed Program. Nature 590, 290–299 (2021).

47. Yang, J., Lee, S. H., Goddard, M. E. & Visscher, P. M. GCTA: A Tool for Genome-wide Complex Trait Analysis. Am J Hum Genet 88, 76–82 (2011).

48. Benjamini, Y. & Hochberg, Y. Controlling The False Discovery Rate - A Practical And Powerful Approach To Multiple Testing. J. Royal Statist. Soc., Series B 57, 289–300 (1995).

49. Mägi, R. & Morris, A. P. GWAMA: software for genome-wide association meta-analysis. BMC Bioinformatics 11, 288 (2010).

50. Smith, S. M. et al. An expanded set of genome-wide association studies of brain imaging phenotypes in UK Biobank. Nat Neurosci 24, 737–745 (2021).

51. Shatokhina, N. et al. ENIGMA-Vis: A Web Portal to Browse, Navigate & Visualize Brain Genome-Wide Association Studies (GWAS). Biological Psychiatry 89, S136 (2021).

52. Seto, M. et al. Multi-Omic Characterization of Brain Changes in the Vascular Endothelial Growth Factor Family during Aging and Alzheimer’s Disease. Neurobiol Aging 126, 25–33 (2023).

53. Leeuw, C. A. de, Mooij, J. M., Heskes, T. & Posthuma, D. MAGMA: Generalized Gene-Set Analysis of GWAS Data. PLOS Computational Biology 11, e1004219 (2015).

54. Elliott, L. T. et al. Genome-wide association studies of brain imaging phenotypes in UK Biobank. Nature 562, 210–216 (2018).

55. Lu, Q. et al. A Powerful Approach to Estimating Annotation-Stratified Genetic Covariance via GWAS Summary Statistics. Am J Hum Genet 101, 939–964 (2017).

56. Zhang, Y. et al. SUPERGNOVA: local genetic correlation analysis reveals heterogeneous etiologic sharing of complex traits. Genome Biology 22, 262 (2021).

57. Gamazon, E. R. et al. A gene-based association method for mapping traits using reference transcriptome data. Nat Genet 47, 1091–1098 (2015).

58. Zorniak, M., Clark, P. A. & Kuo, J. S. Myelin-forming cell-specific cadherin-19 is a marker for minimally infiltrative glioblastoma stem-like cells. J Neurosurg 122, 69–77 (2015).

59. Breitfeld, J. et al. Genetic dissection of serum vaspin highlights its causal role in lipid metabolism. Obesity (Silver Spring) 31, 2862–2874 (2023).

60. Yang, L., Chen, S. J., Yuan, G. Y., Wang, D. & Chen, J. J. Changes and clinical significance of serum vaspin levels in patients with type 2 diabetes. Genet Mol Res 14, 11356–11361 (2015).

61. Zhou, B. et al. Serum Vaspin Levels in Gestational Diabetes Mellitus: A Meta-Analysis. Metab Syndr Relat Disord 21, 535–544 (2023).

62. Goodwin, E. C., Motamedi, N., Lipovsky, A., Fernández-Busnadiego, R. & DiMaio, D. Expression of DNAJB12 or DNAJB14 Causes Coordinate Invasion of the Nucleus by Membranes Associated with a Novel Nuclear Pore Structure. PLoS One 9, e94322 (2014).

63. Akiyama, H., Nakadate, K. & Sakakibara, S. Synaptic localization of the SUMOylation-regulating protease SENP5 in the adult mouse brain. Journal of Comparative Neurology 526, 990–1005 (2018).

64. Yamada, S., Sato, A., Ishihara, N., Akiyama, H. & Sakakibara, S. Drp1 SUMO/deSUMOylation by Senp5 isoforms influences ER tubulation and mitochondrial dynamics to regulate brain development. iScience 24, 103484 (2021).

65. Sarachana, T. & Hu, V. W. Genome-wide identification of transcriptional targets of RORA reveals direct regulation of multiple genes associated with autism spectrum disorder. Mol Autism 4, 14 (2013).

66. Acquaah-Mensah, G. K., Agu, N., Khan, T. & Gardner, A. A Regulatory Role for the Insulin- and BDNF-Linked RORA in the Hippocampus: Implications for Alzheimer’s Disease. Journal of Alzheimer’s Disease 44, 827–838 (2015).

67. Al-Zaid, F. S., Hurley, M. J., Dexter, D. T. & Gillies, G. E. Neuroprotective role for RORA in Parkinson’s disease revealed by analysis of post-mortem brain and a dopaminergic cell line. NPJ Parkinsons Dis 9, 119 (2023).

68. van der Meer, D. et al. Understanding the genetic determinants of the brain with MOSTest. Nat Commun 11, 3512 (2020).

69. Rabinowitz, Y. S., Dong, L. & Wistow, G. Gene expression profile studies of human keratoconus cornea for NEIBank: a novel cornea-expressed gene and the absence of transcripts for aquaporin 5. Invest Ophthalmol Vis Sci 46, 1239–1246 (2005).

70. Liu, Y.-P., Zhang, T.-N., Wen, R., Liu, C.-F. & Yang, N. Role of Posttranslational Modifications of Proteins in Cardiovascular Disease. Oxidative Medicine and Cellular Longevity 2022, 3137329 (2022).

71. Mendler, L., Braun, T. & Müller, S. The Ubiquitin-Like SUMO System and Heart Function: From Development to Disease. Circ Res 118, 132–144 (2016).

72. Kurowska, P. et al. Review: Vaspin (SERPINA12) Expression and Function in Endocrine Cells. Cells 10, 1710 (2021).

73. Zhang, H., Schneider, T., Wheeler-Kingshott, C. A. & Alexander, D. C. NODDI: practical in vivo neurite orientation dispersion and density imaging of the human brain. Neuroimage 61, 1000–16 (2012).

